# Smallpox bioterrorism scenarios and reactive intervention protocol: mathematical model-based analysis

**DOI:** 10.1101/2024.11.22.24317777

**Authors:** Youngsuk Ko, Yubin Seo, Jin Ju Park, Eun Jung Kim, Jong Youn Moon, Tark Kim, Joong Sik Eom, Hong Sang Oh, Arim Kim, Jin Yong Kim, Jacob Lee, Eunok Jung

**Affiliations:** Institute of Mathematical Sciences, Konkuk University, Seoul, Korea; Department of Mathematics, Konkuk University, Seoul, Korea; Division of Infectious Disease, Department of Internal Medicine, Kangnam Sacred Heart Hospital, College of Medicine, Hallym University, Seoul, Korea; National Assembly Research Service, Seoul, Korea; Department of preventive medicine, College of medicine, Gachon University, Incheon, Korea; Division of Infectious Diseases, Soonchunhyang University Bucheon Hospital, Bucheon, Korea; Division of Infectious Diseases, Department of Internal Medicine, Gil Medical Center, College of Medicine, Gachon University, Incheon, Korea; Division of Infectious Disease, Department of Internal Medicine, Hallym University Sacred Heart Hospital, Gyeonggi-do, Korea; Gachon Biomedical Convergence Institute, Gachon, Korea; Division of Infectious Diseases, Department of Internal Medicine, Incheon Medical Center, Incheon, Korea

## Abstract

Smallpox, caused by the variola virus, is one of the most devastating diseases in human history and was eradicated through global vaccination efforts by 1980. Despite its eradication, the virus remains in high-security laboratories for research purposes, posing the potential risk of bioterrorism. This study developed a mathematical model to analyze potential smallpox epidemics by incorporating factors such as age groups, heterogeneous contact patterns, and various intervention strategies including contact tracing, ring vaccination, and mass vaccination. The model simulations indicated that the Republic of Korea’s current plans for negative-pressure isolation beds should suffice under most scenarios, but extreme worst-case scenarios could overwhelm healthcare capacity. This study highlights the critical importance of non-pharmaceutical interventions and strategic vaccination prioritization for controlling outbreaks. These findings provide valuable guidance for public health officials and policymakers in preparing for potential bioterrorism threats and emerging infectious diseases. Furthermore, emphasizes the need for comprehensive preparedness and robust response strategies. The proposed framework applies to smallpox and to other infectious diseases, offering insights for future outbreak management.

## Introduction

Smallpox, caused by the variola virus, is one of the most devastating diseases affecting humans(1). With its origins traced back to ancient civilizations, smallpox has spread across continents. The disease is characterized by a high fever, severe skin eruptions, and a significant fatality rate (approximately 30%), making it a formidable threat to populations worldwide.

The pioneering findings of Jenner and concerted global efforts, including mass vaccination campaigns, surveillance, and implementation of ring vaccination strategies, led to the gradual decline of smallpox cases. Non-pharmaceutical interventions (NPIs), such as disease surveillance, case finding, and contact tracing, are also crucial for containing the spread of the virus(2). Ring vaccination involves vaccinating all individuals near a detected case to prevent the spread of the virus. Additionally, post-exposure prophylaxis (PEP) has been used to vaccinate individuals exposed to the virus, further preventing outbreaks(3,4). The World Health Organization (WHO) launched an intensified eradication program in 1967, which culminated in the declaration of smallpox eradication in 1980.

Despite its eradication, the variola virus remains in two high-security laboratories: the Centers for Disease Control and Prevention in the United States and the State Research Center of Virology and Biotechnology in Russia(5). The virus is retained for research purposes, including developing new vaccines and treatments. However, the presence of these viral stocks poses the potential risk of bioterrorism. Given the lack of widespread immunity in the current global population, the deliberate release of the smallpox virus could lead to a catastrophic outbreak. Two notable laboratory-related smallpox incidents underscored the importance of maintaining vigilance. In 1971, the Aral Smallpox incident in Soviet Russia led to ten infections and three deaths(6). Similarly, in 1978, a laboratory accident in Birmingham, England resulted in two infections and one death(7).

A notable scenario that underscores the potential threat of smallpox as a bioterrorism agent is Dark Winter exercises (8). This senior-level bioterrorist attack simulation depicted a covert smallpox attack in the United States starting in Oklahoma City and rapidly spreading to other states. The exercise revealed significant gaps in the national emergency response, highlighting the challenges of containing the outbreak, managing public panic, and maintaining essential services. Winter’s findings emphasize the need for robust preparedness plans, including sufficient vaccine stockpiles, effective communication strategies, and coordinated efforts between public health and security agencies, to mitigate the impact of such a bioterrorism event.

Mathematical models are crucial in understanding and controlling infectious disease outbreaks, including smallpox, as demonstrated in various studies. Ferguson emphasized the effectiveness of targeted surveillance and containment interventions such as ring vaccination in controlling smallpox outbreaks, underscoring the need for a rapid response (9). Meltzer constructed a model to evaluate quarantine and vaccination interventions after a bioterrorist attack and demonstrated that a combination of these strategies was effective in halting disease transmission (10). Ohushima developed a model to predict smallpox outbreaks in Japan, evaluated the control measures, and found that mass vaccination was more effective than ring vaccination under certain conditions (11). Chun used epidemic modeling and tabletop exercises to prepare public health officials in the ROK for potential outbreaks, highlighting the importance of these tools in estimating cases, deaths, and resource shortages (12).

In this study, we developed a mathematical model to analyze potential smallpox epidemics by incorporating various realistic factors such as age groups and heterogeneous contact patterns. The model includes contact tracing, ring vaccination, and mass vaccination strategies and distinguishes severe cases among infected individuals to discuss the required capacity for severe patients in emergency scenarios. The insights gained from vaccination prioritization during the COVID-19 pandemic were also analyzed in relation to potential smallpox scenarios. Given the threat of smallpox as a bioterrorism agent, this study evaluated reactive intervention protocols by simulating various outbreak scenarios and assessing the effectiveness of NPIs, vaccination strategies, and isolation facilities. These findings highlight the importance of early detection, rapid response, and strategic vaccination prioritization. The proposed framework addresses smallpox and offers insights applicable to other emerging infectious diseases, emphasizing the need for comprehensive preparedness in the face of potential bioterrorism threats.

## Materials and Methods

### Mathematical modeling of smallpox epidemic

To investigate the transmission dynamics of potential smallpox epidemics, we developed a susceptible-infectious-recovered-type mathematical model that reflects contact tracing, disease severity, age group, and ring/mass vaccination. The mathematical model is shown as a flow diagram in Figure 1. Age groups are denoted by subscript (*i*) for each variable, and vaccinated individuals are denoted by superscript (*ν*). In this study, we categorized the population into 16 age groups, each spanning 5 years, ranging from 0–4 years to 65 years and older in the Republic of Korea (13).

**Figure 1.**
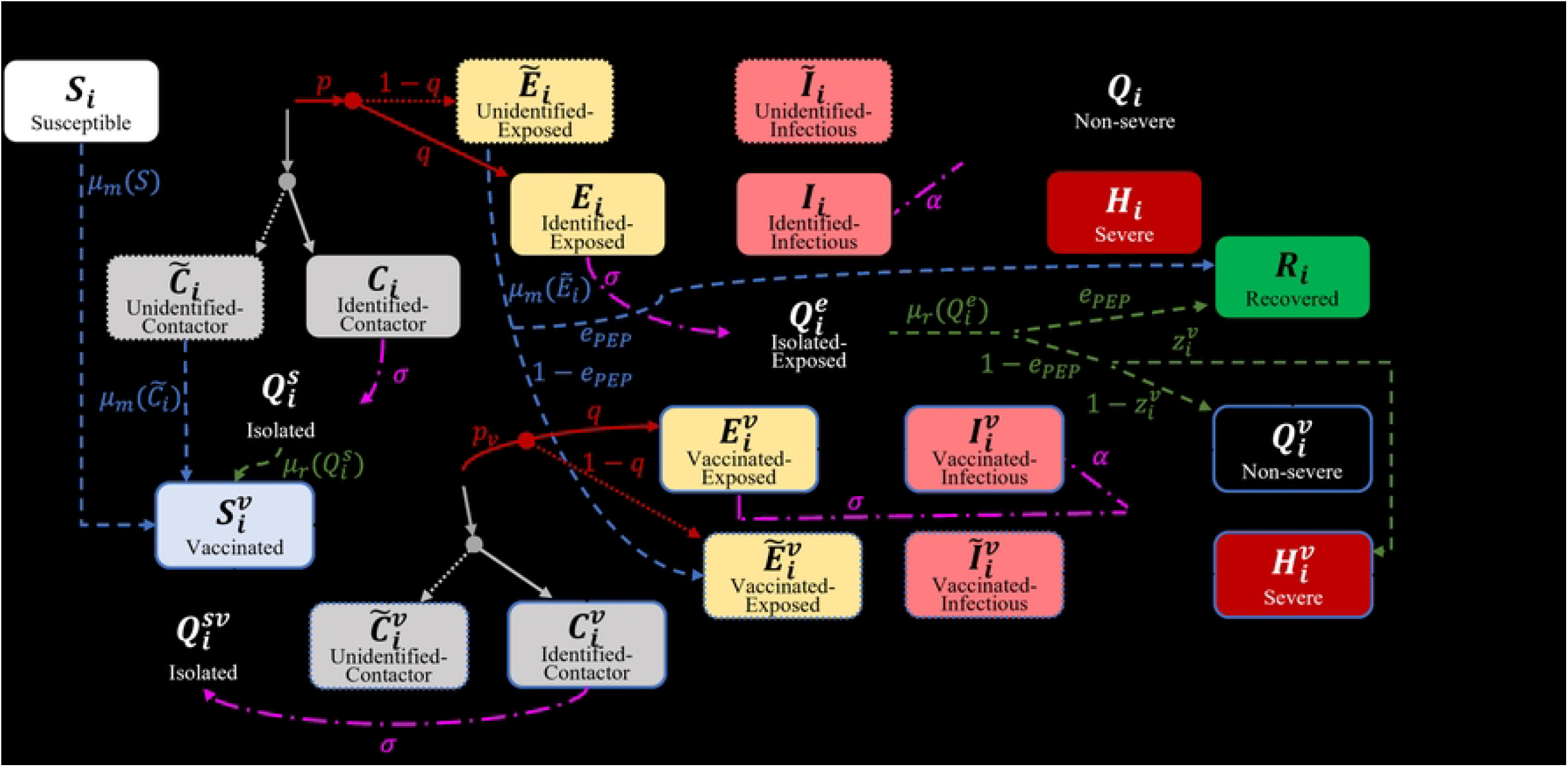
Flow diagram of the mathematical model of the smallpox epidemic.

In general, Susceptible-Infectious-Recovered type models have the transmission rate, typically denoted by the symbol *β*, which consists of the contact rate per unit time (*c*) and the probability of successful disease transmission (*p*), that is, *β* = *pc*. In this study, we distinguished between those who had contact with infectious hosts but were not infected (*C*_*i*_) and those who were infected after contact (*E*_*i*_). We also considered both close contact 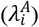and social contact 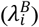. Close contacts were incorporated based on a study by Prem et al.(14). Social contacts were assumed to be four times the number of contacts, excluding household contacts. The probability of successful disease transmission through close contact was set to 60%(15). Using this value and the next-generation matrix method, the basic reproductive number through close contact was calculated to be approximately four (16). Considering that the recorded basic reproductive number of smallpox is approximately six, the probability of successful disease transmission through casual contact was set at 10%(17).

To incorporate contact tracing regardless of infection status, we used the symbol (*q*) to represent the proportion of contact-traced individuals. Those who were traced after contact and those who were not traced were distinguished using a tilde symbol (*∼*). Here, casual contact was not traced. The infection transmission periods for traced and non-traced individuals were set based on the time from symptom onset to isolation during the COVID-19 pandemic in the ROK (18). We assumed that contact-traced individuals would be hospitalized/isolated relatively quickly (within 2.3 days, 1/α) after symptom onset, while non-traced individuals would be isolated after 6.8 days 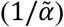, considering the time to the appearance of definitive smallpox symptoms (lesions)(19). The severity rate in the model (*z*_*i*_) was set to twice the infection fatality ratio. Thus, in the model simulations, half of the severe patients died, whereas there were no deaths among non-severe patients.

Vaccination was applied in two forms in the model: mass vaccination (*µ*_*m*_) and ring vaccination (*µ*_*r*_). Ring vaccination was prioritized and administered after case isolation but not in those who had already developed symptoms. Patients exposed to the infection either recovered or continued to show symptoms depending on the effectiveness of PEP (*e*_*PEP*_). Even those who did not directly receive the effects of PEP experienced a reduction in severity/fatality rates due to partial effects. Mass vaccination was administered to the entire population outside the ring vaccination targets, and those who had already been vaccinated were not revaccinated even if they were contact-traced. Those who were isolated without infection or ring vaccination were discharged 19 days (1/*l*) after isolation(20). In response to the smallpox epidemic, we assumed that vaccinating 40 million people (approximately 80% of the population) would be necessary to achieve herd immunity. Vaccination can be slow initially owing to the lack of available personnel who are educated and vaccinated. In the ROK, there was a small-scale vaccination for healthcare workers during the global Mpox outbreak in 2022 in response to the domestic influx (21). These individuals would be the first to start vaccination during a smallpox epidemic and could vaccinate other healthcare workers and target populations. Thus, we assumed the daily vaccination capacity follows a logistic growth model, considering the maximum daily vaccination capacity during the COVID-19 pandemic, set at 1 million (*µ*_*ub*_). The vaccination process started with 1,000 vaccinations per day, with a logistic growth rate (*r*_*ν*_) of 0.1, and all 40 million people were vaccinated after 110 days.

The model is formulated using ordinary differential equations as follows:

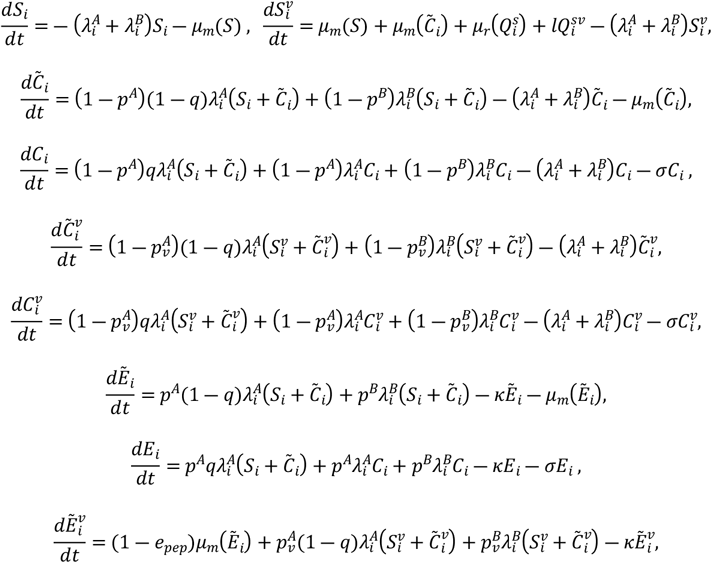

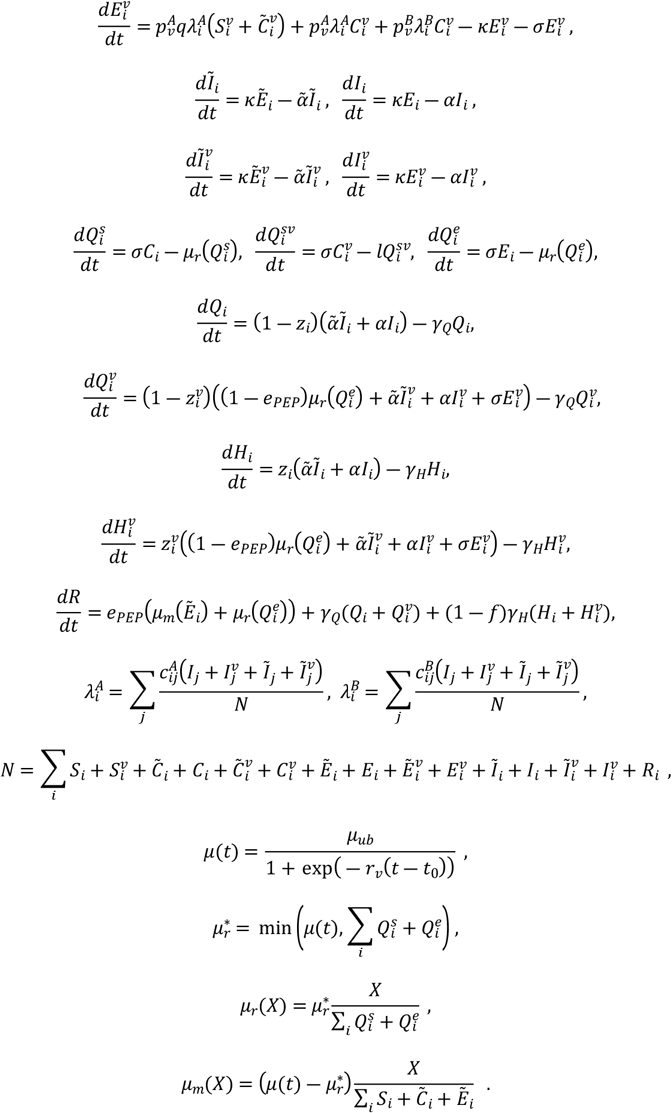

The model parameters are listed in Table 1. As experienced during the COVID-19 pandemic, the scale of the potential outbreaks remains uncertain. To reflect this, model simulations were conducted as a stochastic process using the Tau-leaping method. We ran 1000 simulations for each model setting. As an initial condition for the model simulation, all population groups were assumed to be in a susceptible state. The number of exposed hosts was set to 100 and proportionally distributed according to the population ratio of each group. Table 2 lists the population numbers in each group.

**Table 1.** Model parameters.

| Symbol | Description | Value | Reference |
| --- | --- | --- | --- |
| $1/\tilde{\alpha}$ | Infectious period of contact-unidentified hosts | 6.8 days | (18,19) |
| $1/\alpha$ | Infectious period of contact-identified hosts | 2.3 days | (18) |
| $p_A$ | Probability of successful disease transmission per close contact | 0.6 | (15,16,18) |
| $p_B$ | Probability of successful disease transmission per casual contact | 0.1 | (15,16,18) |
| $q$ | Contact-identification ratio | 0.8 | Assumed |
| $1/l$ | Isolation duration of uninfected case | 19 days | (20) |
| $1/\sigma$ | Contact tracing duration | 2 days | Assumed |
| $1/\kappa$ | Incubation period | 12 days | (22) |
| $1/\gamma_Q$ | Isolation duration of non-severe case | 28 days | (22) |
| $1/\gamma_H$ | Duration from hospitalization to recovery (or death) of severe case | 13 days | (1) |
| $z_i$ | Age dependent severity rate | 0.83 (1)<br>0.47 (2,3)<br>0.61 (4,5)<br>0.59 (6-10)<br>0.64 (11-16) | (23) |
| $f$ | Fatality rate of severe case | 0.5 | Assumed,<br>(1) |
| $e$ | Preventive vaccine effectiveness against infection | 0.78 | (24) |
| $e_H$ | Vaccine effectiveness against severity | 0.97 | (24) |
| $e_{pep}$ | Post-exposure prophylaxis vaccine effectiveness against infection | 0.5 | (25) |

**Table 2.**
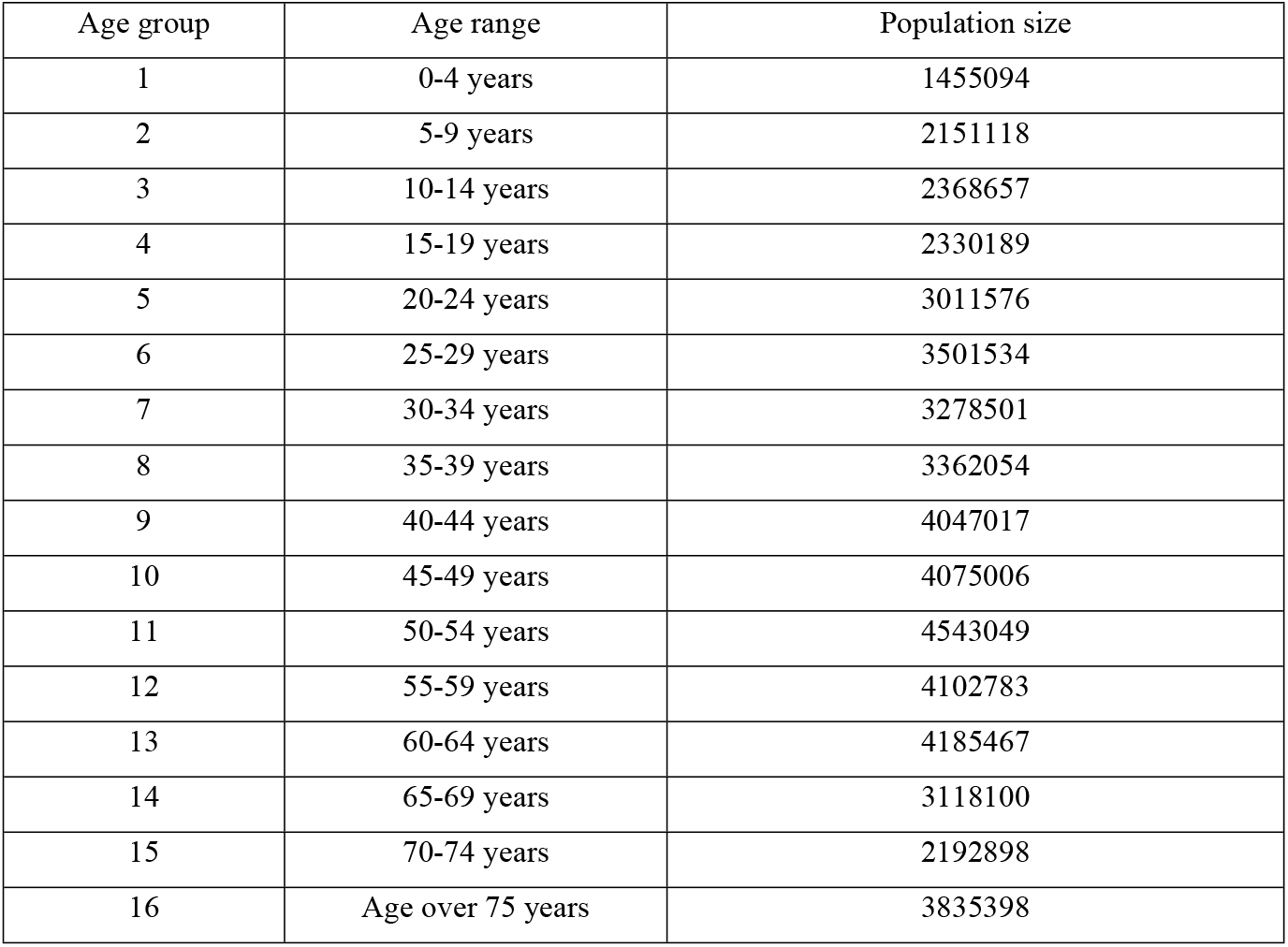
Population size by age group.

| Age group | Age range | Population size |
| --- | --- | --- |
| 1 | 0-4 years | 1455094 |
| 2 | 5-9 years | 2151118 |
| 3 | 10-14 years | 2368657 |
| 4 | 15-19 years | 2330189 |
| 5 | 20-24 years | 3011576 |
| 6 | 25-29 years | 3501534 |
| 7 | 30-34 years | 3278501 |
| 8 | 35-39 years | 3362054 |
| 9 | 40-44 years | 4047017 |
| 10 | 45-49 years | 4075006 |
| 11 | 50-54 years | 4543049 |
| 12 | 55-59 years | 4102783 |
| 13 | 60-64 years | 4185467 |
| 14 | 65-69 years | 3118100 |
| 15 | 70-74 years | 2192898 |
| 16 | Age over 75 years | 3835398 |

### Baseline model simulation scenario

The simulation time was set as 365 days. The baseline model simulation scenario consisted of three phases following the initial exposure:

▸ Pre-declaration (Phase 1): This phase represents the period during which the occurrence of the outbreak has not yet been recognized. No NPIs or vaccination measures were in place during this phase, which lasted for 28 days after the initial exposure. This is the worst-case scenario, set at a realistic level, considering the incubation period, initial symptoms, and occurrence of rashes.
▸ Post-declaration (Phase 2): This phase begins when the outbreak is first recognized, and contact tracing and social distancing (NPIs) are implemented. However, vaccination is not yet feasible because of the time required for preparation, e.g., training for medical personnel. This phase lasted for three days. It was assumed that social distancing would reduce the total contact rate by 60%.
▸ Post-vaccination (Phase 3): This phase marks the beginning of vaccination. Vaccination continued until 40 million people had been vaccinated. This phase lasted for 334 days. Vaccination was administered simultaneously to all age groups, and the amount of vaccines administered was proportional to the population of each group; that is, there was no vaccine prioritization.

### Scenarios considering vaccine prioritization

Vaccination was not prioritized in the baseline scenario considered in this study. However, as experienced by most countries during the COVID-19 pandemic, prioritizing vaccination may be a more realistic approach. We defined four criteria for the prioritization scenarios: ascending age, descending age, prioritizing age groups with higher transmission risk, and prioritizing age groups with higher severity/death risk. The age groups with higher transmission risk were calculated based on the contact rate and the probability of successful disease transmission, and the order was as follows: age groups 4 (15-19 year), 3 (10-14 year), 9 (40-44 year), 8 (35-39 year), 7 (30-34 year), 6 (25-29 year), 10 (45-49 year), 11 (50-54 year), 5 (20-24 year), 2 (5-9 year), 12 (55-59 year), 13 (60-64 year), 1 (0-4 year), 14 (65-69 year), 15 (70-74 year), 16 (75+ year).

## Results

### Baseline scenario simulation

Figure 2 shows the model simulation results for the confirmed cases (transition from infectious to isolated) and isolated patients. The curves represent the mean values of the model simulations, and shaded areas indicate the 95% credible interval (CrI). The red graph shows the daily confirmed cases, whereas the blue curves represent isolated patients. Among the blue curves, the solid and dashed lines indicate patients with non-severe and severe patients.

**Figure 2.**
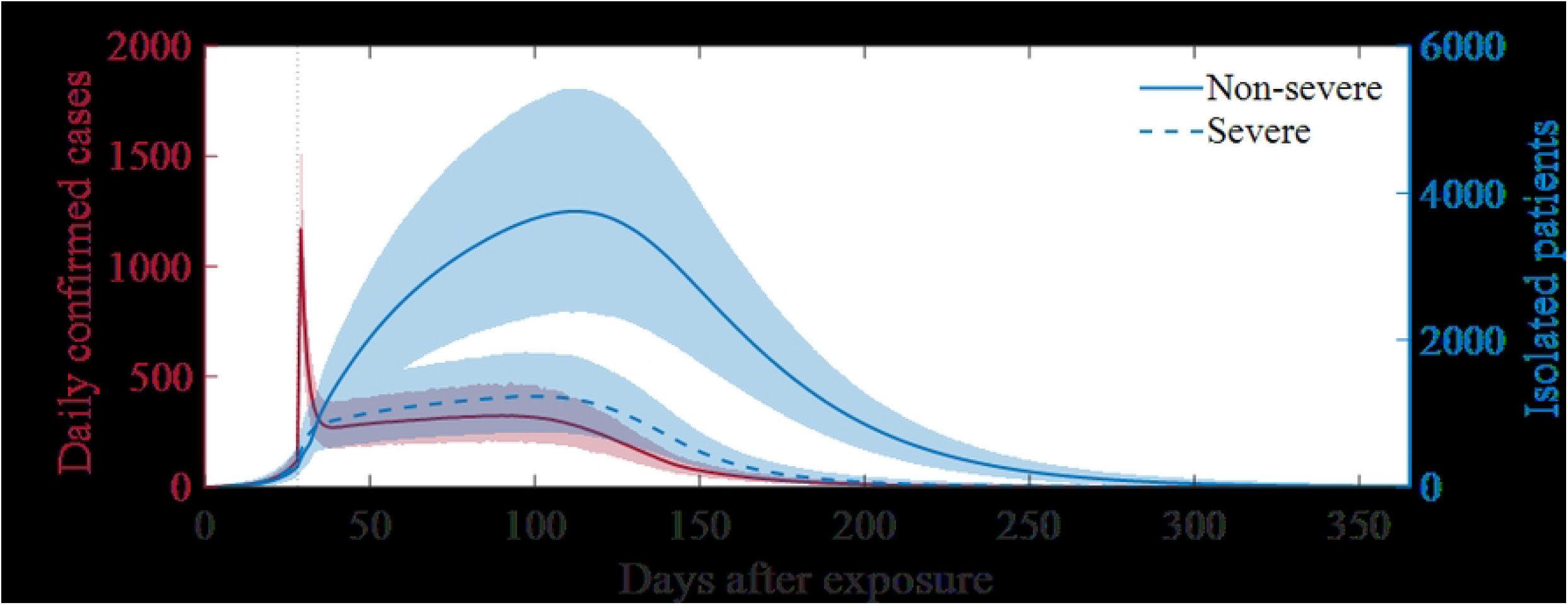
Baseline scenario simulation results. Curves indicate mean simulation results and shaded areas indicate the 95% credible interval.

Daily confirmed cases increased sharply (mean 1167, maximum 1602 in 95% CrI) owing to contact tracing implemented at the initial outbreak recognition, and then decreased, followed by a gradual increase, and finally decreased again owing to herd immunity from vaccination. The number of non-severe patients reached a mean of 3750 (maximum 5422 in 95% CrI) after 114 days of spread, whereas that of severe patients reached a mean of 1235 (maximum 1838 in 95% CrI) after 99 days of exposure.

The confirmed cases and deaths observed in the simulation runs are presented in Figure 3 using box-and-whisker plots. Panels A and B show the confirmed cases, and panels C and D show the number of deaths. The total number of confirmed cases was 36600 (95% CrI [24253, 51500]) and the total number of deaths was 5345 (95% CrI [3472, 7545]). Age group 9 (40–44 years) had the highest number of confirmed cases, with a mean of 4241 (95% CrI [2846, 5969]). Group 11 (50–54 years) had the highest number of deaths, with a mean age of 677 years (95% CrI [447, 936]). The age group 0–4 years had the lowest number of confirmed cases (mean 361, 95% CrI 237, 513]) and deaths (mean 68, 95% CrI [41, 101]).

**Figure 3.**
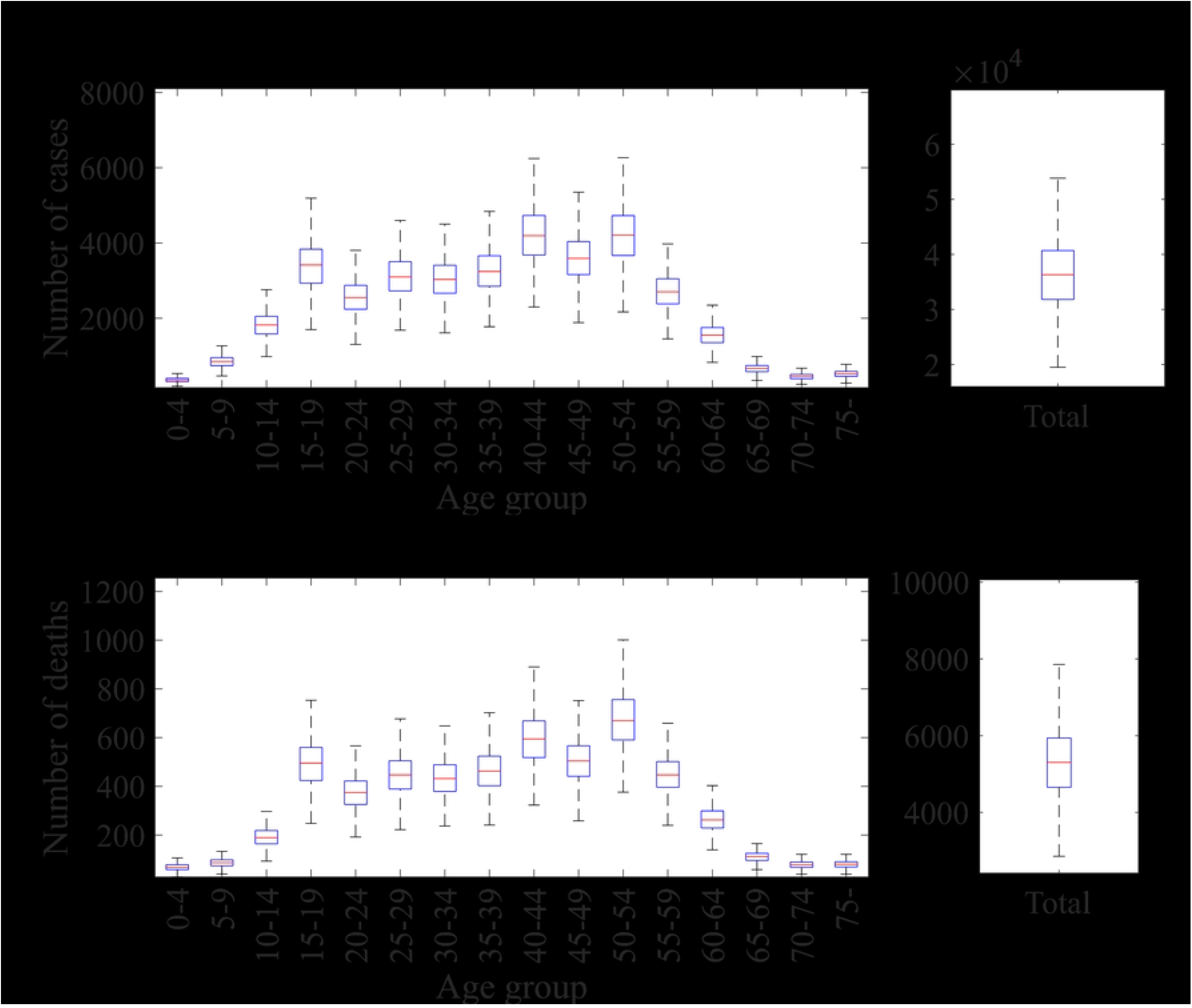
Outbreak outcomes from the baseline scenario simulations: Distribution of the number of confirmed cases by age (A) and total (B), deaths by age (C) and total (D).

In addition to the baseline scenario, the results for different initial numbers and logistic growth rates for vaccinations and the varying effects of social distancing on contact reduction are listed in Tables 3–5.

**Table 3.** Mean and 95% CrI of outbreak outcomes with a 50% reduction in contacts due to social distancing.

| Initial vaccination number | Growth rate | Confirmed cases | Deaths | Peak severe patients |
| --- | --- | --- | --- | --- |
| 1000 (baseline) | 0.05 | 260458 [170462, 354688] | 34255 [22388, 46734] | 7608 [4983, 10423] |
|  | 0.1 (baseline) | 90947 [60018, 127048] | 11890 [7842, 16516] | 3182 [2080, 4452] |
|  | 0.5 | 29323 [19519, 40916] | 3816 [2588, 5298] | 1369 [909, 1915] |
| 5000 | 0.05 | 144457 [94173, 199367] | 19008 [12540, 25960] | 4411 [2875, 6109] |
|  | 0.1 (baseline) | 64585 [41862, 90666] | 8451 [5588, 11793] | 2381 [1562, 3349] |
|  | 0.5 | 26828 [17542, 37368] | 3491 [2316, 4847] | 1282 [849, 1796] |
| 10000 | 0.05 | 111421 [73076, 157301] | 14671 [9640, 20653] | 3507 [2303, 4973] |
|  | 0.1 (baseline) | 56267 [38326, 76475] | 7358 [5030, 9959] | 2135 [1458, 2922] |
|  | 0.5 | 26075 [17039, 37098] | 3391 [2269, 4682] | 1261 [819, 1759] |

**Table 4.** Mean and 95% CrI of outbreak outcomes with a 60% reduction in contacts due to social distancing.

| Initial vaccination number | Growth rate | Confirmed cases | Deaths | Peak severe patients |
| --- | --- | --- | --- | --- |
| 1000 (baseline) | 0.05 | 60356 [40271, 84492] | 8894 [5950, 12436] | 1475 [985, 2094] |
|  | 0.1 (baseline) | 36886 [24398, 51467] | 5391 [3581, 7513] | 1281 [838, 1786] |
|  | 0.5 | 18611 [12524, 25865] | 2676 [1827, 3706] | 1048 [707, 1455] |
| 5000 | 0.05 | 46071 [30001, 63246] | 6784 [4461, 9343] | 1318 [848, 1808] |
|  | 0.1 (baseline) | 30803 [20538, 42278] | 4495 [2996, 6173] | 1208 [797, 1682] |
|  | 0.5 | 17614 [11806, 24104] | 2530 [1713, 3410] | 1028 [691, 1411] |
| 10000 | 0.05 | 40645 [26835, 56652] | 5981 [3918, 8371] | 1261 [827, 1787] |
|  | 0.1 (baseline) | 28100 [19317, 39537] | 4098 [2792, 5702] | 1168 [790, 1642] |
|  | 0.5 | 17103 [11403, 23652] | 2457 [1669, 3365] | 1012 [684, 1404] |

Figure 4 shows the distribution of hosts who had contact with infected individuals at outbreak recognition 28 days after initial exposure. The average numbers of hosts who had contact (whether they were infected or not), hosts during the incubation period, and hosts in the contagious stage were 5618 (95% CrI [3786, 7776]), 3401 (95% CrI [2288, 4706]), and 926 (95% CrI [623, 1275]), respectively.

**Figure 4.**
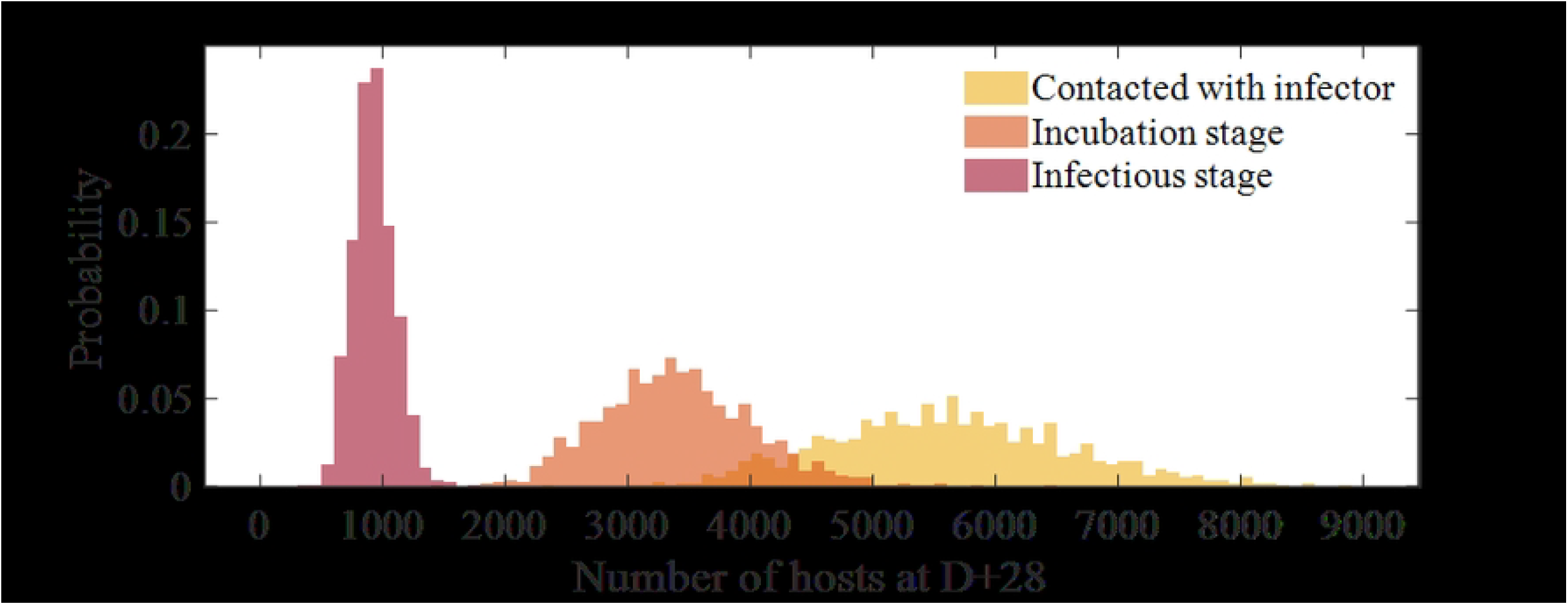
Distribution of the number of hosts who had hazardous contact until the outbreak recognition in different stages.

Figure 5 shows the mean daily vaccination number (panel A), the distribution of administered ring vaccinations in the simulation runs (panel B), and the ring vaccination period (panel C). In this study, the ring vaccination period was considered the point at which the mass vaccination exceeded the amount of ring vaccination. The average number of ring vaccinations administered was 28750 (95% CrI [19145,40218]), and the vaccination period lasted an average of 5.53 days (95% CrI [3.5, 8]).

**Figure 5.**
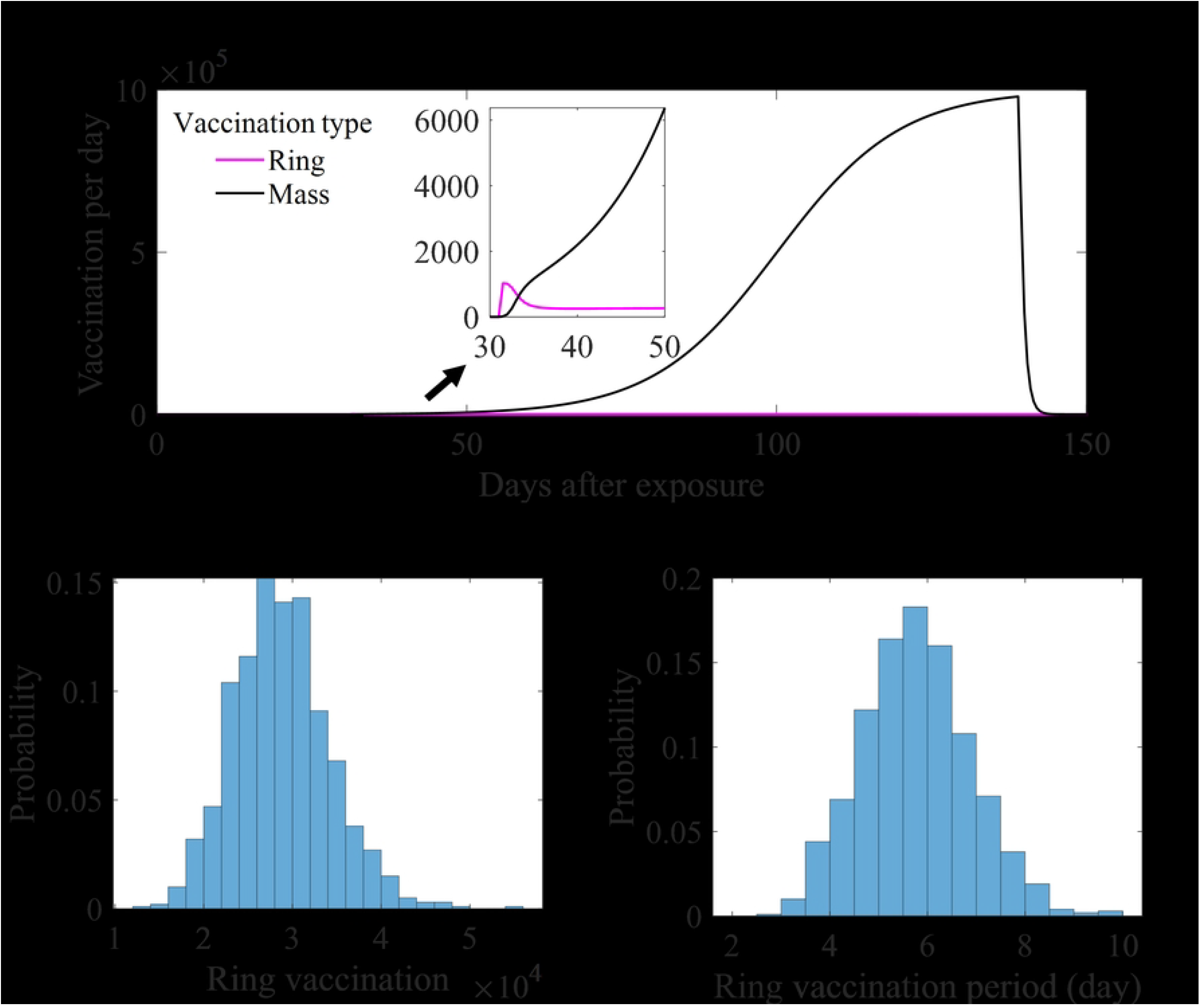
Vaccination number in the baseline scenario simulations: Mean daily vaccination number (A), distribution of administered ring vaccination, and period (B,C).

### Impact of vaccine prioritization

The simulation results, including the baseline scenario and scenarios with vaccine prioritization, are shown in Figure 6. Panels A and B show the daily numbers of confirmed and severe cases, respectively. Compared with the baseline scenario, prioritizing vaccination for age groups with a higher transmission risk (purple) showed a decrease, whereas prioritizing vaccination for older age groups (yellow) showed a significant increase. Table 5 lists the odds ratios for cumulative confirmed cases, deaths, and the peak number of severe patients compared to the baseline scenario. When prioritization based on transmissibility was applied, the odds ratios for all metrics were below one, within the confidence interval. Conversely, when prioritization based on descending age was applied, the odds ratios for all metrics exceeded one.

**Table 5.** Mean and 95% CrI of outbreak outcomes with a 70% reduction in contacts due to social distancing.

| Initial vaccination number | Growth rate | Confirmed cases | Deaths | Peak severe patients |
| --- | --- | --- | --- | --- |
| 1000 (baseline) | 0.05 | 20749 [13713, 29494] | 3370 [2225, 4805] | 894 [602, 1256] |
|  | 0.1 (baseline) | 17565 [11933, 23971] | 2851 [1947, 3852] | 881 [605, 1186] |
|  | 0.5 | 12510 [8588, 17000] | 1985 [1358, 2681] | 890 [611, 1224] |
| 5000 | 0.05 | 19069 [13225, 26800] | 3101 [2105, 4343] | 891 [625, 1237] |
|  | 0.1 (baseline) | 16446 [11093, 22420] | 2663 [1853, 3608] | 893 [596, 1228] |
|  | 0.5 | 12006 [8172, 16475] | 1898 [1321, 2584] | 884 [598, 1215] |
| 10000 | 0.05 | 18288 [12598, 25139] | 2975 [2042, 4084] | 891 [625, 1210] |
|  | 0.1 (baseline) | 15724 [11007, 21809] | 2545 [1781, 3533] | 889 [609, 1224] |
|  | 0.5 | 11900 [8028, 16371] | 1879 [1285, 2577] | 890 [596, 1228] |

**Table 6.**
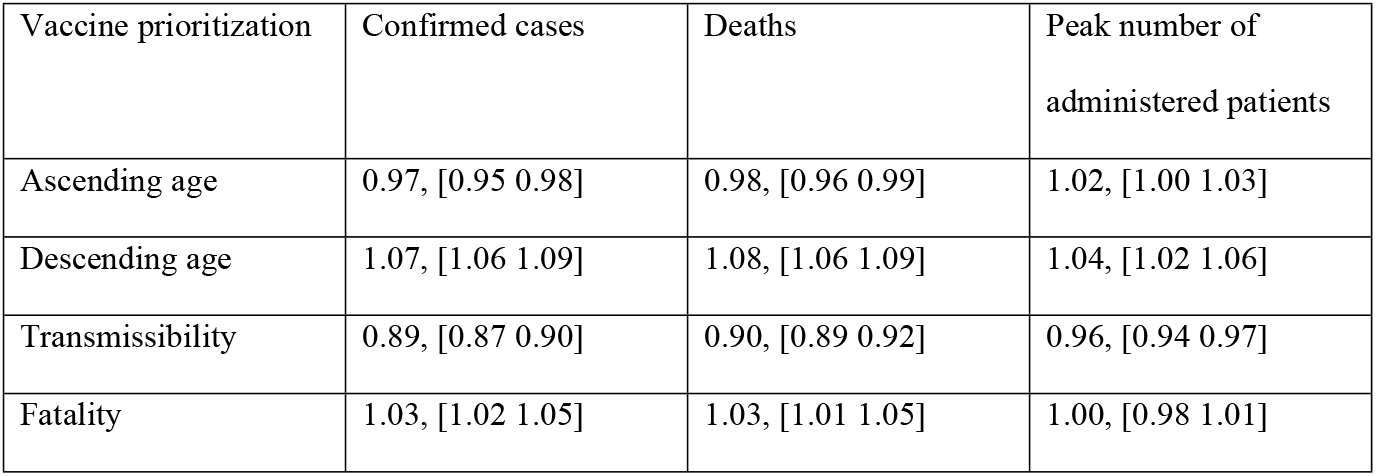
Odds ratios and confidence intervals for scenarios considering vaccine prioritization compared to the baseline scenario.

**Figure 6.**
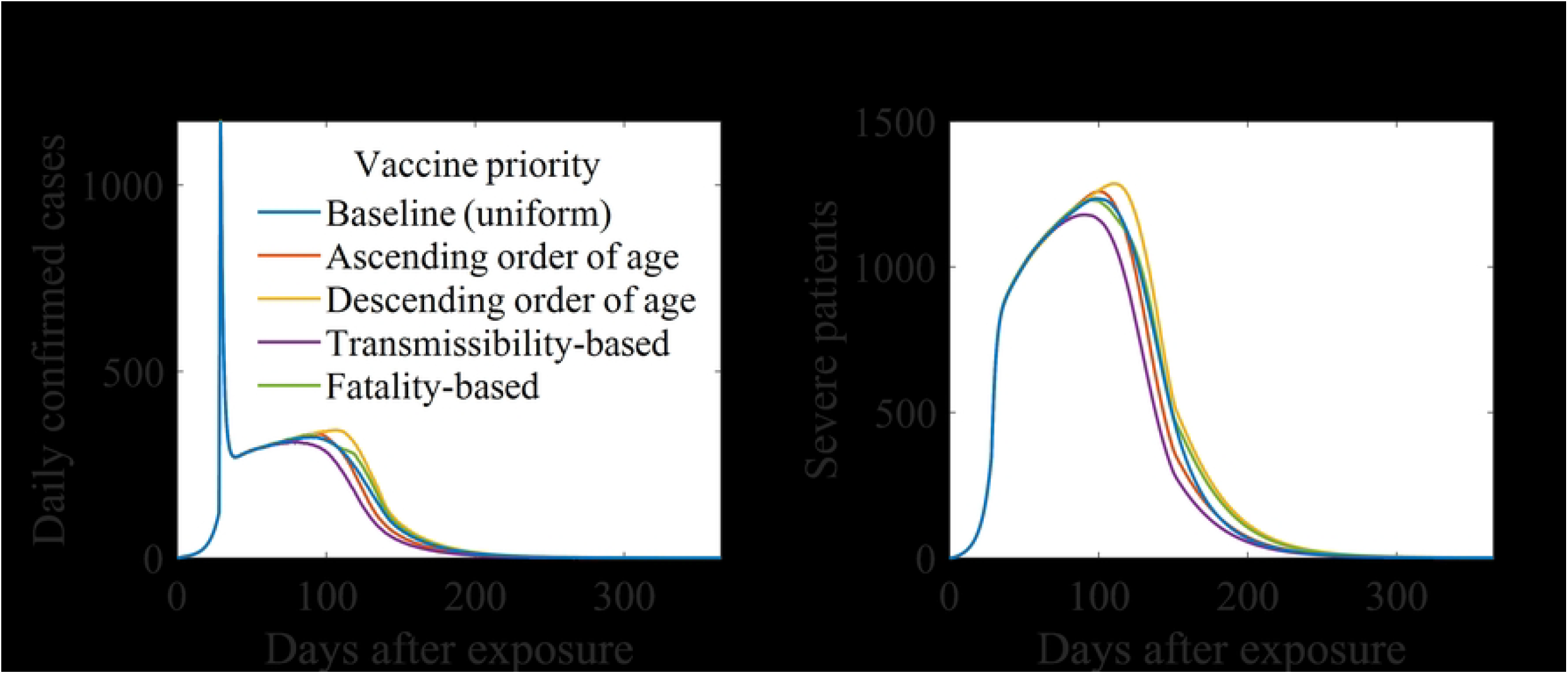
Simulation results considering vaccine prioritization: Daily confirmed cases (A), administered to severe patients (B).

### Sensitivity analysis

To address the inherent uncertainties in these values and conduct a comprehensive sensitivity analysis of the model outcomes, we measured the partial rank correlation coefficient (PRCC) values using Latin hypercube sampling. PRCC is a statistical measure used to determine the strength and direction of the relationship between two variables while controlling for the effects of other variables. In sensitivity analysis, it is particularly useful to identify the parameters that have the most significant impact on the output of a model. A detailed description of this method is provided in reference (26). We considered the following model inputs: outbreak recognition timing, impact of social distancing on contact number, infectious period of traced and non-traced cases, contact identification ratio, and logistic growth rate of the daily vaccination number. The model outputs were set as the cumulative confirmed cases and deaths, and the peak number of administered severe patients.

Figure 7 shows the measured absolute values of PRCC over time. The colors in the graph represent the model inputs. Solid curves indicate the values for cumulative confirmed cases, whereas solid curves indicate the values for cumulative deaths. The order of the absolute PRCC values for the model inputs was the same regardless of whether the model output was cumulative confirmed cases or deaths. Based on the final absolute PRCC values, the timing of outbreak recognition had the highest value, with 0.90 for cumulative confirmed cases and deaths. The contact identification ratio initially had a relatively high absolute PRCC (0.29) for cumulative confirmed cases but decreased to 0.01, resulting in the lowest PRCC. Table 7 lists the ranges of PRCC values. In contrast to Figure 7, the absolute value of the PRCC for the growth rate of daily vaccination was the second smallest when the considered model output peaked for severe patients. Additionally, outbreak recognition timing had the highest absolute PRCC.

**Table 7.** The final value and range of PRCC of model inputs considering different model outputs. There is no range if the target model output is the peak number of severe patients as there is a one-time point of it.

| Model input | Model output |  |  |
| --- | --- | --- | --- |
|  | Cumulative confirmed cases | Cumulative deaths | Peak severe patients |
| Outbreak recognition timing | 0.90 [0, 0.93] | 0.90 [-0.19, 0.93] | 0.88 |
| Impact of social distancing | -0.93 [-0.93, 0.01] | -0.91 [-0.91, 0] | -0.85 |
| Isolation rate for traced cases | -0.28 [-0.28, 0] | -0.32 [-0.32, 0.04] | -0.27 |
| Isolation rate for non-traced cases | -0.89 [-0.89, -0.26] | -0.88 [-0.88, 0.2] | -0.81 |
| Contact-identification ratio | -0.01 [-0.29, -0.01] | -0.01 [-0.01, 0.01] | -0.02 |
| Growth rate of the daily vaccination | -0.31 [-0.31, 0] | -0.33 [-0.33, 0] | -0.16 |

**Figure 7.**
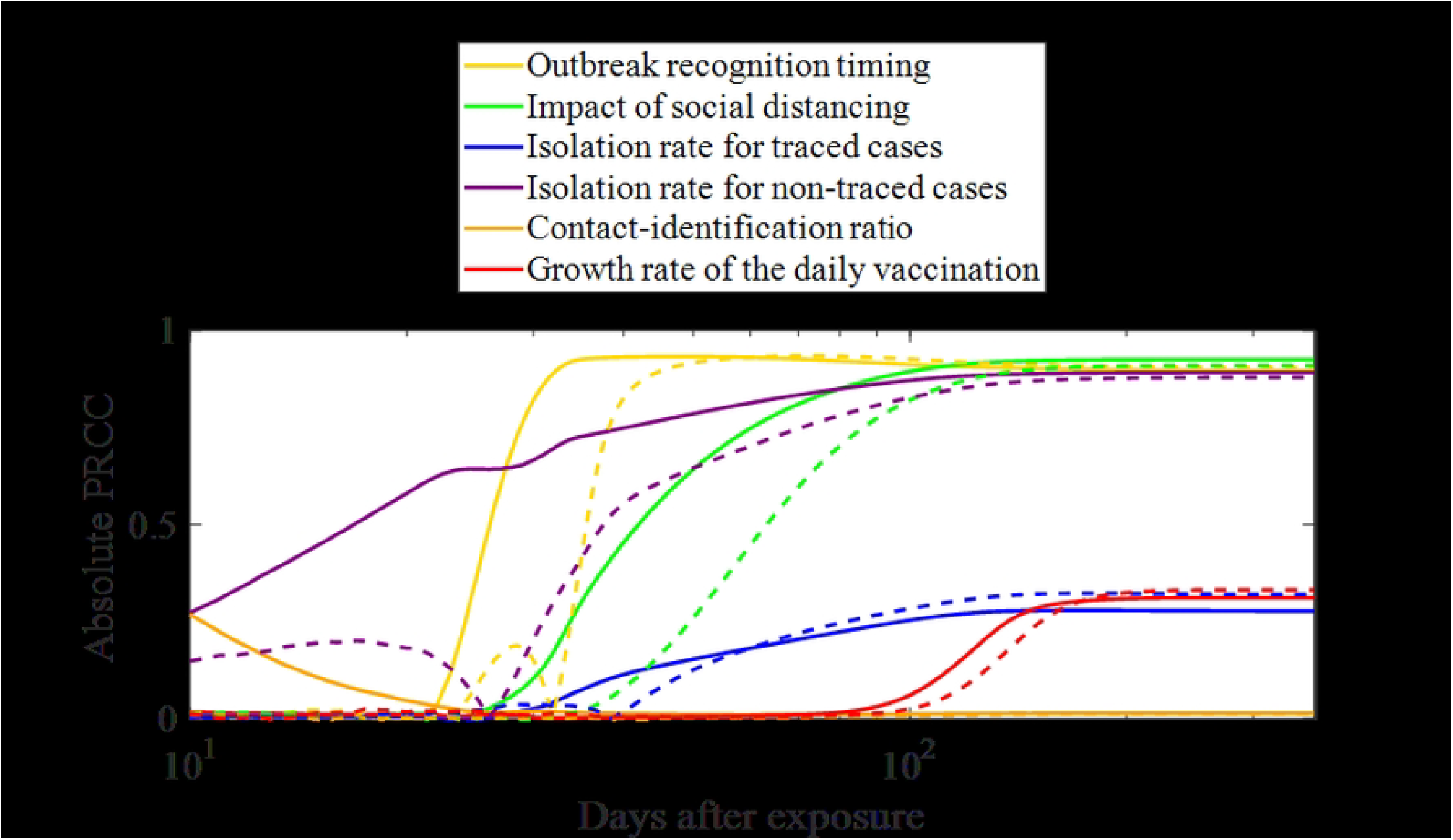
Absolute value of PRCC over time. Solid lines represent the value for cumulative confirmed cases, while dashed lines represent the value for cumulative deaths.

## Discussion

The model simulation showed a rapid increase in confirmed cases upon initial detection, followed by a gradual increase in the number of severely ill patients. By 2024, the ROK plans to expand the number of negative-pressure isolation beds to 3500 to respond to emerging infectious diseases(27). According to the baseline scenario results, even in the worst case within the 95% CrI, the peak administered to severely ill patients was 1800 (Figure 2), indicating that the current plan should prevent a shortage of beds. However, in an extreme worst-case scenario, where the vaccination rate is low and the impact of social distancing is weak (upper row in Table 3), the number of administered severe cases could reach up to 7600, posing a significant risk. However, if the effectiveness of social distancing reaches at least 0.6, dangerous situations are avoided. Therefore, the results suggest that a minimum level of NPIs required during a smallpox outbreak. This level was measured to be near the level of social distancing stage 2 in previous COVID-19 studies and is not an unrealistic measure in a bioterrorism situation where nationwide interventions would be stricter.

The parameter sensitivity analysis results provided additional support for the basic model simulation outcomes. The analysis revealed that both deaths and confirmed cases were highly sensitive to recognition timing, impact of social distancing, and isolation rate for non-traced cases (Figure 7 and Table 7). This underscores the significant role of NPIs in achieving herd immunity through vaccination.

The distribution of hosts with contagious contacts at the initial outbreak recognition (Figure 4) represents the minimum number of ring vaccination targets that need to be considered immediately. Considering the additional real-time increase in the number of contacts and exposed individuals, the distribution shown in Figure 5 indicates the overall scope of the ring vaccination plan. Although these individuals represented a small proportion (approximately 0.1%) of the total vaccination scale, ring vaccination was primarily conducted during the first week of the initial vaccination period. Our findings serve as a basis for determining the operation and number of distribution points during this period.

Determining vaccine priorities is challenging because of various social, economic, and ethical issues. Similar problems have been encountered during the COVID-19 pandemic. Prioritizing vaccination for the elderly, who have higher severity/fatality rates, was the best strategy for minimizing deaths when social distancing measures were in place to reduce the effective reproduction number to approximately one (28,29). However, if the effective reproduction number increases, prioritizing the elderly would be less effective than vaccinating younger adults in minimizing deaths. The study was based on the original strain of COVID-19, which had a basic reproductive number of approximately three, roughly half of that of smallpox. This implies that strong NPIs can effectively suppress the spread. Conversely, for smallpox, for which moderate levels of NPIs were not sufficient to control the spread, prioritizing vaccination for groups with higher transmission rates was more effective in reducing deaths. Prioritizing the elderly yielded the least favorable results. If the vaccination history of the elderly, which was not considered in this study, were accounted for, they would likely have a relatively lower severity/fatality rate compared to other age groups, further diminishing the effectiveness of the first vaccination strategy.

The limitations of this study are as follows. For social contacts, we only used estimates based on close contacts. Age-specific severity rates were derived from data that included both vaccinated and unvaccinated individuals, which may differ from the actual values. We did consider the smallpox vaccinations administered in the ROK until the early 1970s and rather assumed that all population groups were susceptible. However, despite the lack of vaccine effectiveness against infection, the elderly might have a lower severe/fatality rate than other age groups due to their vaccination history. Finally, although the smallpox vaccine can have significant side effects, we did not incorporate these side effects into our model. This was because vaccination coverage was fixed. Future studies will focus on analyzing optimal vaccination strategies, taking into account side effects, spatial heterogeneity, and regional lockdowns.

## Conclusion

Based on the findings of this study and considering realistic intervention scenarios and outbreak situations, we propose an appropriate number of isolation facilities for severely ill patients and the necessary level of initial social distancing. Various simulations have highlighted the critical importance of early detection and rapid responses to mitigate the impact of smallpox outbreaks. These results underscore the need for robust preparedness plans that include vaccination and NPIs.

Our study emphasizes the importance of strategic vaccination prioritization and the role of NPIs in controlling outbreaks. The insights gained from this study provide valuable guidance to public health officials and policymakers in preparing for and responding to potential bioterrorism threats and emerging infectious diseases. The critical importance of early detection, rapid response, and comprehensive preparedness cannot be overstated when safeguarding public health.

The overall framework of this study applies to smallpox and other emerging infectious diseases that may spread to humans in the future. By incorporating parameters similar to those applied in this study, response strategy scenarios can be developed for diseases that can be controlled using currently available vaccines. Conversely, for novel diseases with significant time requirements for vaccine development, this framework can be adapted to simulate the post-declaration phase responses.

## Data Availability

All relevant data are within the manuscript.

## Abbreviations

NPIs: Non-pharmaceutical interventions
PEP: Post-exposure prophylaxis
ROK: Republic of Korea
CrI: Credible interval
PRCC: Partial Rank Correlation Coefficient

## Acknowledgement

This research was supported by the Government-wide R&D Fund Project for Infectious Disease Research (GFID), Republic of Korea (grant No. HG23C1629). This work was supported by the Research Program funded by the Korea Disease Control and Prevention Agency (정책, 150).

## Notes

### Competing Interest Statement

The authors have declared no competing interest.

### Funding Statement

Yes

### Author Declarations

This study is a simulation-based research, with no human subjects involved therefore, IRB approval is not required.

